# Inter-Rater Reliability and Usability of CATHIS core for Homeopathic Intervention Studies

**DOI:** 10.1101/2025.11.11.25339874

**Authors:** Martin Loef, Petra Weiermayer, Katharina Gaertner, Daniel Wrzałko, Abhijit Dutta, Subhranil Saha, J.A.D.S. Lakshani, Stephan Baumgartner, Robbert van Haselen

## Abstract

**Background:** CATHIS core is a streamlined appraisal tool for homeopathic intervention studies focusing on credibility, coherence, and clinical relevance, with risk of bias assessed separately (RoB 2/ROBINS-I). The aim of the research project was to evaluate its inter-rater reliability, feasibility, and face validity.

**Methods:** In a preregistered cross-sectional study, four experienced raters independently applied CATHIS core to 28 trials (22 randomised controlled trials, 6 non-randomised studies on interventions) drawn from reviews on insomnia and hypertension; two external reviewers provided consensus ratings. Inter-rater reliability (IRR) was estimated using percent agreement, Fleiss’ κ, and Gwet’s AC2 (95% CIs). Feasibility was quantified as rating time and consensus time. Associations among the three domains were explored with correlation analyses and sensitivity checks.

**Results:** IRR varied markedly by domain. Credibility showed good agreement (Fleiss’ κ = 0.66, 95% CI 0.57–0.74; AC2 = 0.76, 0.71–0.82). Coherence yielded only poor-to-fair agreement (κ = 0.28, 0.16–0.40; AC2 = 0.41, 0.30–0.51). Clinical relevance was similarly limited (κ = 0.32, 0.23–0.41; AC2 = 0.36, 0.28–0.44). Individual ratings required on average 65.8 minutes, while consensus discussions averaged 17.7 minutes. Correlation analyses indicated heterogeneous and partly overlapping domain signals with limited interpretability. Face-validity responses reflected moderate-to-high acceptance but difficulties in consistent application.

**Conclusion:** CATHIS core produced reproducible ratings for credibility but only fair and operationally fragile agreement for coherence and clinical relevance, alongside non-trivial rating burden. Taken together, the current reliability profile is insufficient for confident use in systematic reviews. Targeted refinement—clarifying definitions, tightening scoring rubrics, and simplifying decision rules—appears warranted to improve reproducibility and feasibility before broader implementation.

## Background

Clinical research on homeopathic interventions comprises several hundred randomized and non-randomized studies, yet conclusions remain unsettled. A recent systematic review of six meta-analyses (1990–2023) reported significant effects of homeopathy versus placebo, with evidence quality rated high for individualized and moderate for non-individualized homeopathy according to GRADE [1]. In contrast, a methods-focused assessment of 99 peer-reviewed studies published since 2011, judged most controlled trials to be at high or unclear risk of bias and identified frequent deficits in adherence, safety reporting, and alignment with homeopathic principles [2]. These observations are consistent with earlier programmatic reviews distinguishing individualized from non-individualized approaches but still underscoring methodological limitations [3, 4].

Together, these findings motivate the need for critical appraisal methods that extend beyond internal validity to additional dimensions of evidence quality that are pertinent to whole medical systems. In this regard, research on complementary and alternative medicine has increasingly adopted a whole systems perspective, emphasizing that interventions such as homeopathy should be evaluated within their theoretical and therapeutic context rather than as isolated modalities. Whole systems research highlights the interdependence of internal, external, and model validity, arguing that study designs must align with the explanatory model of the system under investigation and reflect its complexity and real-world delivery [5]. Building on this multidimensional perspective, efforts have been made to develop comprehensive appraisal frameworks that integrate these validity aspects in the evaluation of homeopathic interventions. This perspective underscores the need for appraisal tools that consider not only bias control but also the paradigm fit and contextual coherence of studies within their therapeutic model.

Several instruments address specific facets of study quality. The Cochrane tools RoB 2 and ROBINS-I provide structured assessments of risk of bias in randomized trials (RCTs) and non-randomized studies of interventions (NRSIs), respectively [6, 7]. The Model Validity of Homeopathic Treatment (MVHT) evaluates concordance with homeopathic principles [8], whereas PRECIS-2 and RITES characterize studies on the efficacy–effectiveness continuum and thus speak to external validity and clinical applicability [9, 10]. These instruments cover complementary but only partially overlapping constructs and use non-commensurate outputs, which can reduce comparability across reviews.

To provide a structured and integrative appraisal tailored to homeopathic intervention trials, the original Critical Appraisal Tool for Homeopathic Intervention Studies (CATHIS) was developed by expert consensus to integrate four domains: risk of bias, credibility, coherence, and clinical relevance [11]. An initial validation study reported variable inter-rater agreement, considerable rating time, and inconsistencies in item interpretation, suggesting that simplification might improve feasibility. Also, CATHIS had not yet been evaluated in validation studies conducted by reviewers independent of their development, leaving uncertainty regarding applicability across different user groups.

Against this background, CATHIS core was conceived as a streamlined version retaining three domains—credibility, coherence, and clinical relevance. Domain 1 (risk of bias) was omitted because the original CATHIS items did not adopt RoB 2/ROBINS-I verbatim, which risked redundancy and conceptual misalignment with the current methodological standard. Within CATHIS core, the three retained domains are defined as follows: *credibility* refers to compliance with good clinical practice and the reliability of data, which is supported when a protocol is prospectively published, ethical approval is obtained, protocol deviations are transparently reported, and pseudonymised data are made available after trial completion. *Coherence* denotes concordance of the tested intervention with the principles, materials, strategies, and tools that define the therapeutic model; this requires an accurate and referenced description of the intervention (what, when, how, by whom), its current evidence base, the specific research question, and all procedures used, thereby helping to ensure that investigators tested what they intended to test (model validity). *Clinical relevance* describes the extent to which findings are meaningful for routine care; key determinants include sample size, choice of comparator, patient and setting comparability, intervention flexibility, and the patient-centred importance of outcomes.

This study evaluates the inter-rater reliability and practical feasibility of the CATHIS core for the retrospective assessment of RCTs and NRSIs on homeopathic interventions by raters with clinical experience in homeopathy and includes exploratory analyses of relationships across the three domains. The results are intended to provide an empirical basis for assessing its suitability for future use in systematic reviews.

## Methods

### Study design and registration

We conducted a cross-sectional inter-rater reliability and feasibility study of CATHIS core applied to RCTs and NRSIs on homeopathy. The protocol was preregistered on the Open Science Framework (osf.io/vdf6u). Reporting follows the GRRAS recommendations for reliability and agreement studies [12] (see supplement 3); item-level details of the instrument and the excel tool are provided in an online data repository.

### Sample selection

The selection of included studies was pragmatically constrained and aligned with an ongoing series of systematic reviews on the comparative effectiveness of homeopathy [13]. Accordingly, studies were drawn from literature searches of two systematic reviews covering the conditions insomnia and hypertension. Up to 30 eligible trials were sampled, including both randomized controlled trials (RCTs) and non-randomized studies of interventions (NRSIs). Publications not in English were translated using publicly available automated translation tools; all reviewers were provided with the original and the English version, without instructions on which to prioritise.

### Raters and calibration

Four reviewers with expertise in homeopathy (three physicians and homeopaths, one veterinarian and homeopath) performed independent ratings. After piloting two studies, two additional external reviewers (one physician/homeopath, one epidemiologist/homeopath) issued a consensus decision, resulting in improved guidance to harmonise interpretation; this guidance was incorporated in a new version of the Excel workbook and applied prospectively. For the remaining studies, the four internal ratings were first completed, after which the two external reviewers jointly determined a final consensus score. In doing so, they calculated the mean of the four ratings and identified the corresponding score range. If the mean did not coincide with a five-point increment, it was rounded to the nearest increment, with the direction of rounding informed by their own judgment. When their assessment clearly lay outside the range of internal ratings, the consensus score was set to the respective boundary value. In exceptional cases (include references) the final score was based directly on the consensus reviewers’ appraisal, with explicit rationale provided.

To generate consensus commentaries, we employed an AI-assisted approach. This approach was chosen to enhance consistency in terminology and efficiently synthesize reviewer remarks across numerous studies. A tailored prompt guided the AI to analyse reviewer comments, extract key themes and divergences, and draft brief statements (maximum 60 words) aligned with CATHIS terminology and criteria (see supplement 3). The resulting statements were reviewed and validated by KG and RvH. One reviewer (SS) was co-author of three included studies [14–16]; therefore, his ratings for these studies were excluded from the consensus decisions to avoid potential bias.

### Rating procedure and data capture

Ratings were completed with the CATHIS-core Excel workbook, which implements the scoring guideline and computes domain and total scores. Raters timed completion in minutes per study; time to reach pairwise consensus was also recorded. User-experience items were collected via a brief Google Forms questionnaire (see supplement 3).

### Outcomes

Primary outcome was inter-rater reliability (IRR) at item and domain level. Secondary outcomes were feasibility (rater burden: time per rating; time to consensus) and associations among domains (credibility, coherence, clinical relevance).

### Sample size and monitoring

We used an adaptive sequential approach per Gwet to target a stable percent agreement (Pa) with error margin ≤20% of Pa [17]. An interim analysis after 15 studies rated by four raters was planned to assess whether precision was sufficient; if not, the number of studies would be increased (up to 30) and/or the number of raters extended (to 5–6). Stopping criteria were predefined (Pa stability, E ≤ 20%·Pa, or infeasibility of further expansion).

### Statistical analysis

Inter-rater reliability (IRR) was estimated from the raw ordinal ratings using Gwet’s AC2 (95% CIs) and Fleiss’ κ. Magnitudes of agreement were interpreted using Altman’s conventional thresholds where [0-0.2], [0.21-0.4], [0.41-0.6], [0.61-0.8], and [0.81-1] indicate poor, fair, moderate, good, and very good agreement, respectively [18]. Cumulative interval membership probabilities (cIMP) were applied to derive overall qualitative categories [17].

As prespecified in the protocol, correlations between the three CATHIS core domains—credibility (D1: 0–30), coherence (D2: 0–30), and clinical relevance (D3: 0–50)—were examined on the basis of consensus summary scores. Pearson’s correlation coefficient (r) was used to capture linear associations and Spearman’s rank-order correlation (ρ) to capture monotonic associations.

Additional exploratory sensitivity analyses were performed to assess robustness. These included normalization of domain scores to percentages of their theoretical maxima with descriptive checks for floor/ceiling effects, partial correlations to examine the relationships among domains independent of overall score variance (i.e., controlling for the total score), and non-parametric bootstrap resampling (5,000 iterations) to estimate bias and standard errors, with bias-corrected and accelerated CIs when ceiling effects were suspected.

Finally, after completing their ratings, reviewers also participated in an exploratory face-validity survey, evaluating the clarity, relevance, and usability of CATHIS core with a standardized questionnaire.

All analyses were conducted in R 4.5.1 using standard packages (e.g., irrCAC, psych, polycor, boot). Estimates are reported with CIs; null-hypothesis significance testing was not applied to IRR metrics.

## Results

### Sample size determination

At the interim analysis (15 studies, 4 raters), the predefined precision criterion was already satisfied. Nevertheless, we extended sampling to 28 studies to improve robustness and ensure representation across different clinical indications.

### Study characteristics

In total, 30 records initially met the inclusion criteria (see supplement 1), identified through the literature searches of two systematic reviews (on insomnia and hypertension). Two were subsequently excluded after closer examination: one was found to be a non-unique record of an already included study, and another did not provide original study data. Thus, 28 unique studies were retained for analysis. Of these, 22 were randomized controlled trials (RCTs) and 6 were non-randomized studies of interventions (NRSIs). Twelve studies investigated insomnia, and 16 addressed hypertension. Nineteen studies evaluated a single homeopathic preparation, while eight assessed complex formulations (unclear in one study). Twelve studies examined individualized homeopathy, whereas 15 investigated non-individualized approaches (unclear in one study). Twenty-two studies appeared as journal articles, while six were available in other formats (conference abstracts, theses). Twenty-one studies were published in English, and seven in other languages. The consensus ratings of the 28 studies are provided in the online repository (see data availability statement).

### Interrater reliability

Interrater reliability results are presented in Table 1. For credibility, percent agreement was 0.91, with Fleiss’ κ of 0.66 and Gwet’s AC2 of 0.76; cIMP indicated an Altman classification of good for both, κAC2. Coherence showed lower agreement (Pa = 0.76), with Fleiss’ κ of 0.28 and AC2 of 0.41, corresponding to poor (κ) and fair (AC2). For clinical relevance, Pa was 0.76, Fleiss’ κ 0.32 and AC2 0.36, both classified as fair according to cIMP.

**Tab. 1:**
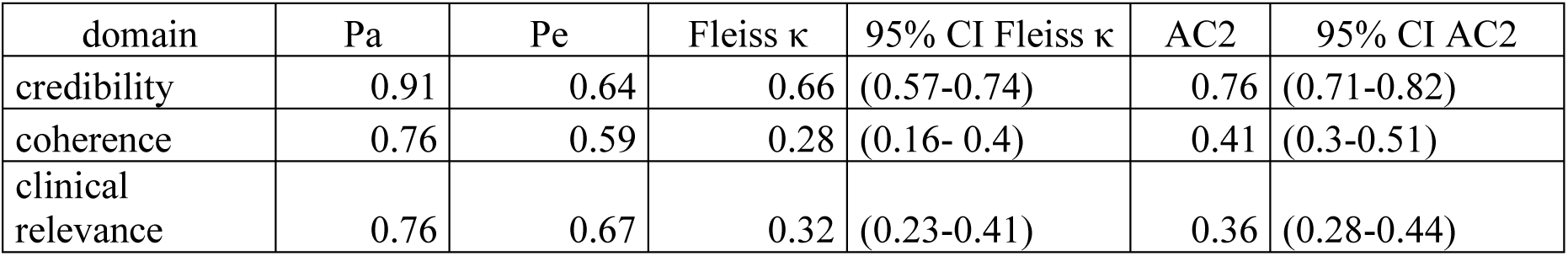
Interrater reliability of CATHIS core domains: percent agreement (Pa), expected agreement (Pe), Fleiss’ Kappa, and Gwet’s AC2 with 95% confidence intervals.

Sensitivity analyses supported the robustness of the main findings (see supplement S2). Excluding studies in which a rater was also a co-author resulted in only minor changes to agreement estimates for all domains. Likewise, reclassification of “not applicable” responses with adjusted weighting did not alter the reliability pattern.

### Usability

Across the included studies, the time required for individual reviewer evaluations (pooled across all four reviewers) averaged 65.8 minutes (SD = 47.5), with a median of 50 minutes (range 2–263; IQR = 45.3; n = 26 studies). The consensus process required substantially less time, with a mean of 17.7 minutes (SD = 11.3), a median of 15 minutes, and durations ranging from 5 to 45 minutes (IQR = 15.0; n = 15 studies).

### Correlation analysis

Consensus summary scores ranged from 0–25 (M = 13.39, SD ≈ 6.1) for credibility (D1), from 0–30 (M

= 21.07, SD ≈ 7.0) for coherence (D2), and from 0–50 (M = 27.68, SD ≈ 9.8) for clinical relevance (D3). The domains differed in their maximum possible scores (30, 30 and 50, respectively), reflecting distinct scales.

#### Bivariate correlations

Pearson and Spearman correlations indicated positive associations among the domains. Credibility showed modest correlations with coherence and clinical relevance, whereas coherence and clinical relevance were closely related. Pearson correlations were r = .32 (p=0.097) for credibility–coherence, r = .54 (p=0.003) for credibility–clinical relevance, and r = .83 (p<0.001) for coherence–clinical relevance. The strong correlation between coherence and clinical relevance may reflect some overlap between model alignment and perceived clinical applicability. Spearman ρ showed a similar pattern, with ρ = 0.19 (p=0.32), ρ= 0.51 (p=0.005) and ρ= 0.71 (p < 0.001) for the respective domain pairs.

#### Partial correlations

When controlling for the total score to account for shared variance across domains, the pattern of associations changed, suggesting that the bivariate correlations were partly driven by overall score level. A strong negative correlation emerged between credibility and coherence (Pearson r = –0.73 (p < 0.001); Spearman ρ = –0.58 (p=0.001)), indicating that, after adjusting for overall score, higher credibility ratings were associated with lower coherence ratings and vice versa. Credibility and clinical relevance were also negatively associated, though to a lesser extent (Pearson r = –0.47 (p=0.014); Spearman ρ = –0.32 (p=0.11)). In contrast, coherence and clinical relevance showed no meaningful partial association (Pearson r =–0.06 (p=0.76); Spearman ρ=0 (p=0.99)). Because each domain contributes to the total score, these partial correlations reflect the relationships between domains after removing the variance they share with the overall quality score.

#### Bootstrap analyses

Nonparametric bootstrapping confirmed the robustness of these findings. Bias-corrected estimates closely matched the observed values, with small biases (0.01-0.02) across all coefficients. Standard errors were largest for the weaker correlations (SE≈0.15–0.19 for credibility with coherence/clinical relevance) and smallest for the coherence–clinical relevance correlation (SE=0.08), consistent with its high stability.

### Face validity

The results of the reviewer questionnaire are reported in figure 1. Exploratory face-validity ratings indicated moderate to high acceptance of CATHIS core. Mean ratings across reviewers ranged from 6.3 (range 3–9) for reproducibility to 9.0 (range 7–10) for novelty. Reviewers judged the instrument as partly challenging to apply, particularly regarding clarity of terms and instructions (means ≈ 6–7). Overall, these results support the perceived novelty and completeness of CATHIS, while suggesting that its clarity, applicability, and reproducibility could be further optimized.

**Fig. 1:**
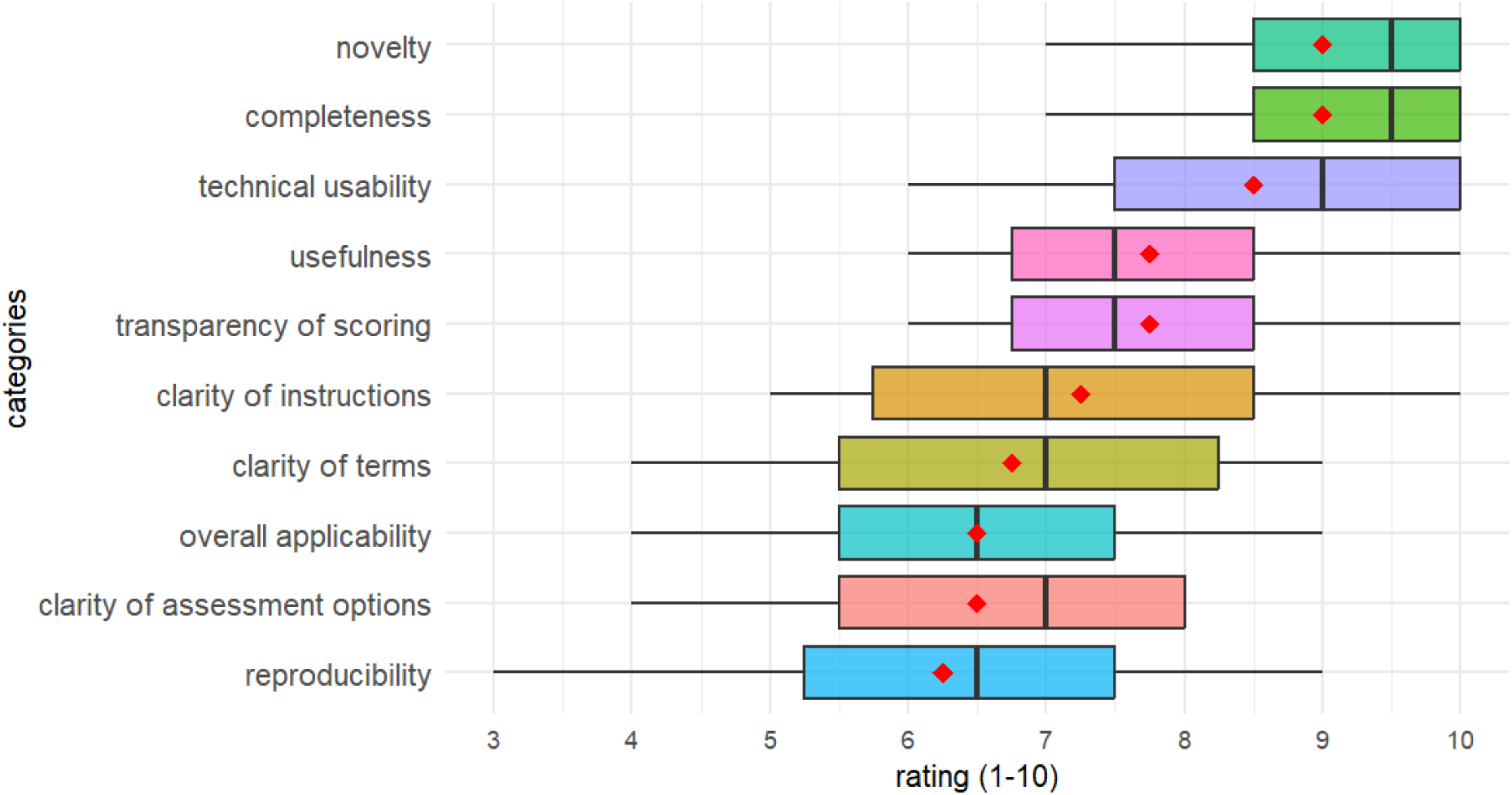
Exploratory face-validity ratings of the CATHIS core instrument (red diamonds=mean, black line is median, box = interquartile ranges, whiskers = absolute range (Min-Max))

## Discussion

This study examined the inter-rater reliability, feasibility, and face validity of CATHIS core in a set of randomized and non-randomized trials of homeopathic interventions. The findings provide the first empirical evidence on the instrument’s performance under routine appraisal conditions.

Inter-rater reliability differed substantially between domains. Credibility achieved good agreement across raters, suggesting that items linked to established standards of clinical research (CONSORT elements such as for instance protocol registration or ethical approval) were comparatively straightforward to assess. In contrast, coherence and clinical relevance showed only poor-to-fair agreement, indicating considerable variability in how raters interpreted and applied the corresponding items. This reduced consistency may stem from greater subjectivity in judging model validity and clinical applicability, as well as from ambiguities in the CATHIS core guidance and items. The results therefore suggest that while credibility can be rated reproducibly, further clarification and standardization are required to improve the reliability of coherence and clinical relevance.

Comparisons with established instruments support our recommendation to refrain from routine use of CATHIS core at this stage. For RoB 2, overall agreement was poor (κ = 0.16) with an average of ∼28 minutes per outcome [19], and for ROBINS-I, agreement on the AC1 scale was substantial (≈ 0.67) but assessments were multi-hour even after familiarization [20]. A reanalysis of the same RoB 2 dataset using Gwet’s AC2 indicated moderate overall agreement (AC2 ≈ 0.59, 95% CI 0.51–0.68), highlighting the sensitivity of reliability estimates to the choice of metric [21]. While such comparisons suggest that limited inter-rater reliability and non-trivial burden are general features of complex appraisal, they are not sufficient, on their own, to recommend CATHIS core for routine application. During the consensus process, several methodological challenges became apparent. One problem was ambiguity in the interpretation, making it possible for experts to ‘overthink’ the meaning of items resulting in multiple possibilities with differing score implications. Also, during consensus discussions it appeared that the rating of model validity and clinical relevance was often influenced by the reporting quality as reflected in the credibility rating, leading to difficult decisions about the extent to which this should be discounted. Some CATHIS items required more detailed specification, particularly regarding their application to different study designs or types of homeopathic interventions. Moreover, reviewers often provided limited written justifications or omitted overall study comments, making it difficult to verify consistent application of the tool. Both RoB 2 and ROBINS-I are supported by comprehensive manuals, signalling-question algorithms, and community calibration resources, whereas CATHIS core currently lacks equivalent scaffolding. Given that our data show reproducibility only for credibility and low agreement for coherence and clinical relevance—precisely where guidance is sparse—further specification, detailed guidance, and calibration exercises should precede wider use.

Bivariate analyses suggested that credibility, coherence, and clinical relevance were positively associated, with particularly strong overlap between coherence and clinical relevance. However, once overall study quality was controlled for, these associations largely disappeared or even reversed. This indicates that the raw correlations mainly reflect a general tendency for higher-rated studies to score well across all domains, rather than specific relationships between the domains themselves. The negative partial association between credibility and coherence may reflect rater inconsistency, methodological artefacts, or genuine divergence between the constructs; available data do not allow a clear distinction.

One possible explanation for the strong negative partial correlation between credibility and coherence is a tension between conventional trial methodology and homeopathic practice principles. Studies adhering closely to clinical research standards may constrain individualization and holistic case-taking, thereby reducing coherence, whereas studies emphasizing homeopathic coherence may sacrifice methodological rigor. Alternatively, the inverse association may reflect rater contrast effects or statistical artefacts from controlling for the overall score. Further work with independent ratings and clearer domain definitions is needed to clarify these possibilities. Bootstrap analyses confirmed the robustness of these patterns. Overall, the correlation results highlight uncertainties in how consistently the domains can be applied and interpreted.

Face-validity ratings indicated moderate acceptance of the instrument, while the substantial time required for individual assessments but shorter consensus discussions underscore the burden of application as well as the potential value of refined guidance.

Our study has several limitations. The sample was drawn pragmatically from two systematic reviews (insomnia, hypertension), which may have introduced a learning effect among raters and thus underestimated the average time per assessment. Thematic homogeneity may also have inflated agreement: one study on AMSTAR IRR found that reliability heavily depended on the specific pair of reviewers, suggesting possible overestimation when raters share similar expertise or focus areas (Pieper et al., 2017). Thus, the reported IRR values may underestimate the variability that could occur with a more heterogeneous, randomly selected set of studies. By contrast, results on face validity are less likely to have been affected, as they primarily reflect the clarity and comprehensibility of the instrument. Further limitations include the relatively homogeneous background of raters, which may have constrained the range of perspectives; the use of machine translations for non-English trials, which could have introduced minor interpretation errors; and the reliance on self-reported timing data, which may not fully reflect the burden under routine review conditions. Finally, because the instrument was adapted during the study, the present findings pertain to a specific iteration of CATHIS core and may not generalize to future versions.

In its further development, CATHIS or CATHIS core will need to address several challenges, including clearer definitions for coherence and clinical relevance, more detailed guidance, and opportunities for rater calibration. Improved applicability and reproducibility are closely linked to the conceptual clarity of the items. Clearer and less overlapping operational definitions may help reduce rater variability and improve consistency across settings. Evaluating RCTs and NRSIs together presented challenges to the raters during the study. Moreover, although a single summary score can simplify quality assessment and synthesis [22], the use of summary scores is debated because they apply arbitrary weightings, may conflate reporting with methodological quality, and often lack construct validity [23]. This can obscure domain-specific strengths and weaknesses of trials, creates spurious precision, and may increase noise due to misclassification when threshold-based judgments are applied.

Beyond these scoring issues, the appraisal of complex constructs such as model validity and external validity is inherently difficult. External validity remains a diffuse construct that is hard to operationalize; no validated, widely accepted instrument for its assessment currently exists [24]. A 2024 Delphi study proposes a definition and a set of candidate criteria, but tool development and validation are still required before routine use [25].

The assessment of validity in complex interventions entails a tension between acknowledging contextual and epistemological dimensions, which resist reductionist approaches [26], and the methodological requirement for reproducible appraisal in systematic reviews. Structured tools such as the Model Validity of Homeopathic Treatment (MVHT) [8] and the RITES instrument for positioning trials on the efficacy–effectiveness spectrum [10] offer structured means to operationalize model and external validity, thereby supporting review conclusions that are both methodologically rigorous and clinically meaningful. Because MVHT and RITES address different constructs and yield non-commensurate metrics, a qualitative integration—articulating how model validity and design pragmatism jointly condition the interpretation of effect estimates—offers a more defensible approach than forcing a level of quantitative comparability that the instruments were not designed to provide.

## Conclusion

Taken together, our findings indicate that CATHIS core, in its present form, provides reproducible ratings only for credibility, while coherence and clinical relevance show limited reliability and considerable rating burden. The instrument therefore requires further refinement before it can be recommended for systematic review use. In the meantime, established instruments such as MVHT and RITES may offer a more practical basis for structured appraisal, particularly when combined through transparent narrative synthesis. Future development and empirical testing will be required to determine its potential role in systematic review methodology.

## Further information

### Support

The study was financially supported by the Software AG-Stiftung, Darmstadt, Germany. The funder played no role in the design, methodology, administration or writing of the study.

### Declaration of generatizve AI and AI-assisted technologies in the writing process

During the preparation of this work the authors used ChatGPT (OpenAI, San Francisco, CA, USA; GPT-5, 2025) in order to improve the clarity and readability of the manuscript text and to refine formatting of R code. After using this tool, the authors reviewed and edited the content as needed and take full responsibility for the content of the published article.

### Data Availability Statement

The data will be made publicly available after peer review and publication of the final manuscript.

## List of Abbreviations

AC2: Gwet’s Agreement Coefficient 2.
AMSTAR: A MeaSurement Tool to Assess Systematic Reviews
CATHIS: Critical Appraisal Tool for Homeopathic Intervention Studies; original tool assessing four domains: risk of bias, credibility, coherence, and clinical relevance.
CATHIS core: Streamlined version of CATHIS retaining three domains: credibility, coherence, and clinical relevance.
cIMP: Cumulative Interval Membership Probabilities;.
CI: Confidence Interval.
CONSORT: Consolidated Standards of Reporting Trials
D1 / D2 / D3: Domains of the CATHIS core instrument: D1 = Credibility; D2 = Coherence; D3 = Clinical Relevance.
GRRAS: Guidelines for Reporting Reliability and Agreement Studies
IRR: Inter-Rater Reliability
IQR: Interquartile Range
MVHT: Model Validity of Homeopathic Treatment.
NRSI: Non-Randomized Study of Interventions
Pa: Percent Agreement
Pe: Expected Agreement
RCT: Randomized Controlled Trial.
RITES: Rating of Included Trials on the Efficacy–Effectiveness Spectrum
RoB 2: Revised Cochrane Risk of Bias Tool for Randomized Trials.
ROBINS-I: Risk of Bias in Non-randomized Studies – of Interventions.
SD: Standard Deviation
SE: Standard Error

## Supplement

### Supplement 1

#### Included studies

1. Bagadia LS, More A. Chapter 4: Stress-induced Anger and Hypertension: An Evaluation of the Effects of Homeopathic Treatment. In: Ovuga E, editor. Stress-Related Disorders. Rijeka: IntechOpen; 2022. p. 79–108.
2. Bavuma N. The effect of a homeopathic complex (Amylenum nitrosum 6CH, Crataegus oxyacantha 6CH, Natrum muriaticum 6CH and Scutellaria lateriflora 6CH) on blood pressure in adults with refractory hypertension. Johannesburg, South Africa: University of Johannesburg; 2016.
3. Bell IR, Howerter A, Jackson N, Aickin M, Baldwin CM, Bootzin RR. Effects of homeopathic medicines on polysomnographic sleep of young adults with histories of coffee-related insomnia. Sleep Medicine. 2011;12(5):505–11. doi: https://doi.org/10.1016/j.sleep.2010.03.013.
4. Bignamini M, Bertoli A, Consolandi AM, Dovera N, Saruggia M, Taino S, et al. Controlled double-blind trial with Baryta carbonica 15 CH versus placebo in a group of hypertensive subjects confined to bed in two old people’s homes. Br Homeopath J. 1987;76(03):114–9. doi: 10.1016/S0007-0785(87)80055-8.
5. Blanco YT, Vernet, M. L., Campo, B. B., Rosales, R. D. &, de Oca MCM. Tratamiento homeopático en pacientes hipertensos de preoperatorio de urgencia. Revista Información Científica. 2011;70(2):18.
6. Carlini EA, Braz, S., & Landfranco, R. P. Efeito hipnótico de medicacao homeopática e do placebo. Avaliacao pela técnica de “duplo-cego” e “cruzamento”. Rev Ass Med Brasil. 1987;33:118.
7. Colombo M Terapia omotossicologica dell’insonnia in età pediatrica. Valutazione di uno studio multicentrico controllato. La medicina biologica. 2003;Oct-Dec.
8. Dutta S, Ganguly S, Mukherjee SK, Ghosh P, Hazra P, Roy AS, et al. Efficacy of individualized homeopathic medicines in intervening with the progression of pre-hypertension to hypertension: A double-blind, randomized, placebo-controlled trial. Explore (NY). 2022;18(3):279–86. Epub 20210605. doi: 10.1016/j.explore.2021.05.007. PubMed PMID: 34147344.
9. Harrison CC, Solomon EM, Pellow J. The effect of a homeopathic complex on psychophysiological onset insomnia in males: a randomized pilot study. Alternative Therapies in Health & Medicine. 2013;19(5):38–43. PubMed PMID: 107968045.
10. Hejazi S, Hosseini Tehrany SA, Salehi Sormaghi MH, Sharifi M. P-1348 - The effects of herbal medicin and homeopathic remedy on insomnia. European Psychiatry. 2012;27:1. doi: https://doi.org/10.1016/S0924-9338(12)75515-3.
11. Hitzenberger G, Rehak PH. Zur Wirkung eines homöopathischen Fertigarzneimittels auf den Blutdruck von Hypertonikern: Eine randomisierte doppelblinde kontrollierte Parallelgruppen-Vergleichsstudie. Wiener Medizinische Wochenschrift. 2005;155(17):392–6. doi: 10.1007/s10354-005-0204-2.
12. Hitzenberger GK, A.; Dorcsi, M.; Bauer, P.; Wohlzogen, F. Kontrollierte randomisierte doppelblinde Studie zum Vergleich einer Behandlung von Patienten mit essentieller Hypertonie mit homöopathischen und pharmakologisch wirksamen Medikamenten. Wiener Medizinische Wochenschrift. 1982;94(24):665–70.
13. Jong MC, Ilyenko L, Kholodova I, Verwer C, Burkart J, Weber S, et al. A Comparative Randomized Controlled Clinical Trial on the Effectiveness, Safety, and Tolerability of a Homeopathic Medicinal Product in Children with Sleep Disorders and Restlessness. Evidence-Based Complementary and Alternative Medicine. 2016;2016(1):9539030. doi: https://doi.org/10.1155/2016/9539030.
14. Kohler R. The treatment of primary hypertension in adult male patients using a homeopathic complex [Master’s dissertation]. Johannesburg, South Africa: University of Johannesburg; 2003.
15. Kolia-Adam N. The efficacy of Coffea Cruda 200cH on insomia. Johannesburg, South Africa: University of Johannesburg; 2010.
16. Master FJ. A study of homœopathic drugs in essential hypertension. Br Homeopath J. 1987;76(03):120–1. doi: 10.1016/S0007-0785(87)80056-X.
17. Michael J, Singh S, Sadhukhan S, Nath A, Kundu N, Magotra N, et al. Efficacy of individualized homeopathic treatment of insomnia: Double-blind, randomized, placebo-controlled clinical trial. Complement Ther Med. 2019;43:53–9. Epub 20190109. doi: 10.1016/j.ctim.2019.01.007. PubMed PMID: 30935555.
18. Mogapi PM. The efficacy of a homoeopathic complex (Crataegus oxycantha 6CH, Viscum album 6CH and Digitalis 6CH) on black adults with essential hypertension. Johannesburg, South Africa: University of Johannesburg; 2014.
19. Montero GD, Valladares MB, Tornes CYLF, Agramonte REA, Calderón JB, Fundora HR. Tratamiento homeopático y convencional de la hipertensión arterial. Revista Médica de Homeopatía. 2016;9(2):53–8. doi: https://doi.org/10.1016/j.homeo.2016.07.005.
20. Naudé DF, Stephanie Couchman IM, Maharaj A. Chronic primary insomnia: efficacy of homeopathic simillimum. Homeopathy. 2010;99(1):63–8. doi: 10.1016/j.homp.2009.11.001. PubMed PMID: 20129178.
21. Prosper C, Rodríguez Sánchez, V. Z., Páez Ochoa, Y., Rodríguez Hernández, R., & García Aguilar, G. Efectividad del tratamiento homeopático en pacientes con trastorno del sueño. Medisan. 2016;20(8):1078–83.
22. Prajakta U. A Clinical Study on The Efficacy of Homoeopathic Medicines in The Treatment of Primary Insomnia – A Pilot Study. African Journal of Biomedical Research. 2024;27:1210–6. doi: 10.53555/AJBR.v27i3.3098.
23. Ronander G. The relative effectiveness of olea europa subsp. africana aqeous on mild to moderate hypertension. Durbon, South Africa: Technikon Natal; 2000.
24. Sadhukhan S, Singh S, Michael J, Misra P, Parewa M, Nath A, et al. Individualized Homeopathic Medicines in Stage I Essential Hypertension: A Double-Blind, Randomized, Placebo-Controlled Pilot Trial. J Altern Complement Med. 2021;27(6):515–21. Epub 20210323. doi: 10.1089/acm.2020.0222. PubMed PMID: 33760643.
25. Saha S, Datta A, Ghosh S, Hossain S, Koley M, Mundle M, et al. Individualized homoeopathy versus placebo in essential hypertension: A double-blind randomized controlled trial. Indian Journal of Research in Homoeopathy. 2013;7:62. doi: 10.4103/0974-7168.116629.
26. Soto, B., Momplet, P., Diaz, P., Perez, R., Gutierrez, M., & Salazar, L. (2020). Tratamiento homeopático y convencional de la urgencia hipertensiva. *Acta Médica del Centro*, *14*, 30–43.
27. Varanasi R, Kolli R, Rai Y, Dubashi R, Reddy GRC, Patole T, et al. Effects of Individualized Homeopathic Intervention in Stage I Essential Hypertension: A Single-Blind Randomized, Placebo-Controlled Trial. Homeopathy. 2020;109(01):A032. doi: 10.1055/s-0040-1702091.
28. Waldschütz R, Klein P. The Homeopathic Preparation Neurexan® vs. Valerian for the Treatment of Insomnia: An Observational Study. The Scientific World Journal. 2008;8(1):689282. doi: https://doi.org/10.1100/tsw.2008.61.

#### Excluded studies

1. Daryani D, Pattanaik N. The Hypotensive Effect of Homoeopathic Medicines In The Management Of The Patients Suffering From Uncomplicated Essential Hypertension - A Prospective, Observational Study. International Journal Of Medical Science And Clinical Invention. 2016. doi: 10.18535/ijmsci/v3i7.06.
2. Flores A HO. Uso de la homeopatia como coadyuvante en el manejo de la hipertension arterial de leve a moderada en pacientes adultos en consulta externa del hospital. Bogota, Colombia: Universidad nacional de Colombia, Faculdad de medicina, 2009.

### Supplement 2

**Tab. S1:** Sensitivity analyses of interrater reliability excluding studies where a rater was co-author and reclassifying “not applicable” responses with adjusted weighting (percent agreement (Pa), expected agreement (Pe), Fleiss’ Kappa, and Gwet’s AC2 with 95% confidence intervals).

| Moderator | domain | Pa | Pe | Fleiss κ | 95% CI Fleiss κ | AC2 | 95% CI AC2 |
| --- | --- | --- | --- | --- | --- | --- | --- |
| Excluding rater's own studies | credibility | 0.92 | 0.61 | 0.61 | (0.51-0.72) | 0.78 | (0.72- 0.84) |
| Reclassification of 'not applicable' responses | credibility | 0.8 | 0.44 | 0.6 | (0.51-0.69) | 0.65 | (0.58- 0.71) |
| Excluding rater's own studies | coherence | 0.75 | 0.61 | 0.27 | (0.15-0.4) | 0.36 | (0.24-0.47) |
| Excluding rater's own studies | clinical relevance | 0.75 | 0.63 | 0.31 | (0.22-0.41) | 0.34 | (0.25- 0.43) |

### Supplement 3

#### Prompt for Chat-GPT to conclude overall reasonings for domains

TASK:

You are an epidemiologist. Your task is to support in finalizing consensus commentaries for each domain in the CATHIS core assessment (D1 Credibility, D2 Coherence, D3 Clinical Relevance) for several studies.

- The file “CATHIS_core_Details.docx” defines:

- the scoring system and point allocation for each domain
- detailed criteria for assessing each domain
- terminology, definitions, and examples of how to justify scores and deductions

Your assignment:

- For each study and each domain (D1, D2, D3):

- Read the individual reviewer comments.
- Identify overlapping arguments, key themes, and differences.
- Write a concise, scientific consensus commentary (max. 60 words in total for all domains) that:

- reflects the consensus score
- summarizes the core points from the individual reviewer comments
- uses terminology and definitions consistent with the guidance in “CATHIS_core_Details.docx”

Example format:

D1: Multiple items not fully covered (e.g. no details on randomisation process, allocation, blinding, or study protocol) and multiple items missing (e.g. sample size, baseline, safety, study registration). D2: missing information on intervention timing, responsible persons, references, and repertorisation tool. D3: eligibility criteria clear, the sample size calculation is missing.

Additional instructions:

- Keep your commentaries concise and strictly scientific.
- Do not invent information beyond what is provided in the reviewer comments or the official documents.

Here are the comments by the raters:

…

Here is a text of the consensus raters regarding these comments:

…

Here is the consensus rating for D1, D2, D3 and total:

…

#### Face validity questionnaire

CATHIS core - Face Validity Survey

**Instructions:**

Please indicate your assessment for each question using a 10-point scale, where **0 represents the least favorable** and **10 the most favorable** evaluation.

1. Clarity of Terms How would you assess the clarity of terms (e.g. *coherence*)?
2. Clarity of Instructions How would you assess the clarity of the instructions and guidance provided?
3. Clarity of Assessment Options How would you assess the clarity of the assessment options (e.g., when to choose “covered”, “some concerns”, or “not covered”)?
4. Overall Applicability How would you assess the overall applicability of the instrument?
5. Novelty To what extent does the instrument add to existing tools assessing study quality?
6. Usefulness How would you assess the overall usefulness of the instrument?
7. Perceived Completeness Does the instrument appear complete in terms of the aspects it considers for evaluating study quality?
8. Technical Usability How would you assess the ease of using the Excel version, including dropdown menus, clickable elements, and automatic scoring?
9. Perceived Reproducibility Do you think other assessors would arrive at similar results when using the instrument as instructed?
10. Perceived Transparency Does the structure and logic of the instrument clearly explain how judgments and scores are derived?
11. Further Comments (optional)

#### GRASS checklist

| Section | Checklist Item | Addressed |
| --- | --- | --- |
| Title and abstract | Identify in title or abstract that interrater/intrarater reliability or agreement was investigated | yes |
| Introduction | Name and describe the diagnostic or measurement device of interest explicitly | yes |
| Introduction | Specify the subject population of interest | yes |
| Introduction | Specify the rater population of interest (if applicable) | yes |
| Introduction | Describe what is already known about reliability and agreement and provide a rationale for the study (if applicable) | yes |
| Methods | Explain how the sample size was chosen. State the determined number of raters, subjects/objects, and replicate observations | yes |
| Methods | Describe the sampling method | yes |
| Methods | Describe the measurement/rating process (e.g., time interval between repeated measurements, availability of clinical information, blinding) | yes |
| Methods | State whether measurements/ratings were conducted independently | yes |
| Methods | Describe the statistical analysis | yes |
| Results | State the actual number of raters and subjects/objects that were included and the number of replicate observations that were conducted | yes |
| Results | Describe the sample characteristics of raters and subjects (e.g., training, experience) | yes |
| Results | Report estimates of reliability and agreement including measures of statistical uncertainty | yes |
| Discussion | Discuss the practical relevance of results | yes |
| Auxiliary material | Provide detailed results if possible (e.g., online) | yes |

